# Lead replacement due to non-infectious reasons in patients undergoing traditional cardiac resynchronization therapy in long-term follow-up

**DOI:** 10.1101/2025.11.17.25340448

**Authors:** Jędrzejczyk-Patej Ewa, Mazurek Michał, Sokal Adam, Kotalczyk Agnieszka, Gumprecht Jakub, Kowalczyk Jacek, Pruszkowska Patrycja, Kowalski Oskar, Lenarczyk Radosław, Kalarus Zbigniew

**Affiliations:** 1^st^ Department of Cardiology and Angiology, Silesian Centre for Heart Diseases, Zabrze, Poland; Department of Human Nutrition, Department of Dietetics, Faculty of Health Sciences, Bytom, Poland, Medical University of Silesia, Katowice, Poland; Division of Medical Sciences in Zabrze, Medical University of Silesia, Katowice, Poland, Department of Cardiology, Silesian Center for Heart Diseases, Zabrze, Poland

**Keywords:** cardiac resynchronization therapy, heart failure, lead, lead replacement, outcome

## Abstract

**Background:** Traditional cardiac resynchronization therapy (CRT) is an effective treatment for patients with heart failure.

**Objective:** The study aimed to determine the long-term durability of CRT leads, factors predisposing to replacement, and the outcomes of lead replacement in CRT patients.

**Methods:** The study population consisted of 1059 consecutive patients with conventional CRT implanted between 2002 and 2019 in a university hospital.

**Results:** During the median follow-up of 1661 days (IQR: 815–2792), a total of 324 leads were replaced in 251 patients (23.7%) for non-infectious reasons. Of those who required lead replacement, 126 subjects (50.2%) had the procedure within the first year since CRT implantation. In patients followed over ten years (n=143; 13.5%), 67 subjects (46.8%) had one or more lead replacements. Duration of index CRT procedure and fluoroscopy time were independent predictors for lead replacement in a multivariable analysis (HR 1.03, 95% CI 1.01–1.06, P=0.02 and HR 1.11, 95% CI 1.02–1.15, P=0.01, respectively). Leads that required replacement were left ventricular (11.2%), right ventricular (10.8%), and right atrial (4%), respectively. As compared with no need for re-intervention, lead-related re-do procedures did not influence overall long-term mortality (55.3% vs 51%, P=0.3) nor incidence of device-related infective endocarditis (5.6% vs 4.5%, P=0.82).

**Conclusions:** Almost 25% of patients with CRT require lead replacement within 4.5 years due to non-infectious reasons, and half of those need replacement within the first year. In ten years, nearly every second patient requires a lead-related re-do procedure. Lead replacement does not affect long-term all-cause mortality.

## Introduction

Traditional cardiac resynchronization therapy (CRT) is an effective method of treatment in patients with symptomatic heart failure (HF) with reduced left ventricular ejection fraction (LVEF) and broadened QRS complex (1, 2, 3). This therapy was first introduced for clinical use in 1996. Many studies have shown that CRT improves quality of life, left ventricle remodeling, functional capacity, and survival. Traditional CRT has advanced technologically over time, with the development of improved left ventricular (LV) leads, along with quadripolar electrodes. The number of patients treated with CRT has risen sharply in recent years (4). Nonetheless, as a complex device with three leads, CRT implantation is inevitably associated with an increased risk of electrode damage, dislocation, dysfunction, or device- or lead-related infection. Previously published data have assessed lead failure rates and the type of failure, but focus on dysfunction in a specific type of lead and do not provide data on the entire population of patients with different types of leads observed over many years.

In addition, the predicted lifespan of HF patients (including CRT recipients) has increased significantly due to recent developments in pharmacotherapy (5), further increasing the likelihood of lead damage and the need for re-do procedure and lead replacement over time. As little is known about the incidence of lead replacement in CRT devices in a very long-term follow-up, we aimed to determine the durability of CRT leads, factors predisposing to their replacement, and long-term outcomes in CRT recipients.

## Methods

### Study population

All consecutive patients with traditional CRT implanted between 2002 and 2019 in a high-volume, tertiary care university hospital in a densely inhabited urban region of Poland were included in the study. Subjects were qualified for CRT implantation in line with the appropriate ESC guidelines, including LVEF ≤ 35%, HF symptoms in New York Heart Association (NYHA) class II, III, or ambulatory class IV (despite optimal medical treatment), and broad QRS complexes. Each patient was informed about the procedure and potential complications and signed an informed written consent. The study complied with international standards, i.e., the Declaration of Helsinki. The study protocol has been approved by the Ethics Committee of the Medical University of Silesia (the Institutional Review Board [IRB] number assigned: BNW/NWN/0052/KB/30/24).

The conventional CRT implantation procedure was performed in accordance with current standards. The placement of the LV lead depended on the venous branch anatomy; the lateral or posterolateral tributary of the coronary sinus was preferred. Before the procedure, all subjects received an intravenous prophylactic antibiotic (cefazolin, single-dose IV, or clindamycin, single-dose IV, in case of cephalosporin allergy).

### Follow-up

All patients were assessed during scheduled (one week, one month, and every six months afterwards) and unscheduled visits throughout the observation period. Hospital records, outpatient notes, telephone calls, insurers’ records, and death certificates directly from patients and relatives were also used for data collection. Patients were followed from CRT implantation until March 2021.

### Statistical analysis

The categorical variables were expressed as numbers and percentages, whereas numerical parameters were expressed as the median with interquartile range (IQR), according to the distribution of the parameters. The groups were compared using the Chi-square, Yates corrected Chi-square, T-Student, or Mann-Whitney U tests as appropriate. Survival was analyzed using the Kaplan-Meier estimator. Multivariable Cox regression tests were used to identify independent risk factors for mortality. The multivariable regression model was constructed to assess the predictors of mortality and included baseline confounders that differentiated alive and deceased patients in the studied population, with a P value of < 0.05, except for redundant variables. Results were expressed as hazard ratio (HR) with 95 percent confidence intervals (95%CI).

The P value of less than 0.05 was considered statistically significant. All statistical analyses were performed using the Statistica software package (version 10.0, StatSoft Inc., Tulsa, OK, USA).

## Results

### Study population

The study population consisted of 1059 consecutive patients with traditional CRT implanted between 2002 and 2019 in a tertiary care university hospital located in a densely populated, urban region of Poland (949 subjects [89.6%] with CRT-D and 110 patients [10.4%] with CRT-P). The median LVEF in the whole population was 25% (IQR: 20-29%), with a median age of 65 years (58–72), 78.6% of males (n=832), and a median QRS duration was 163 msec (150–181).

During the median follow-up of 1661 days (IQR: 815–2792) for non-infectious reasons (dislocation, dysfunction, fracture, etc.), 324 leads in 251 patients (23.7%) were replaced. Sixty patients (5.7%) had 2 lead replacement procedures, 12 subjects (1.1%) had 3 procedures, and 2 of them (0.2%) had 4 lead-related procedures. Of 324 lead replacements, the reasons for re-do procedures were as follows: 67 (20.7%) lead dislocations, 78 (24.1%) lead fractures, 79 (24.4%) high stimulation thresholds, 44 (13.6%) phrenic nerve stimulation and inability to reprogram the device, and 56 (17.2%) low lead impedance. The cumulative incidence of lead replacement has been illustrated in Figure 1A.

**Figure 1.**
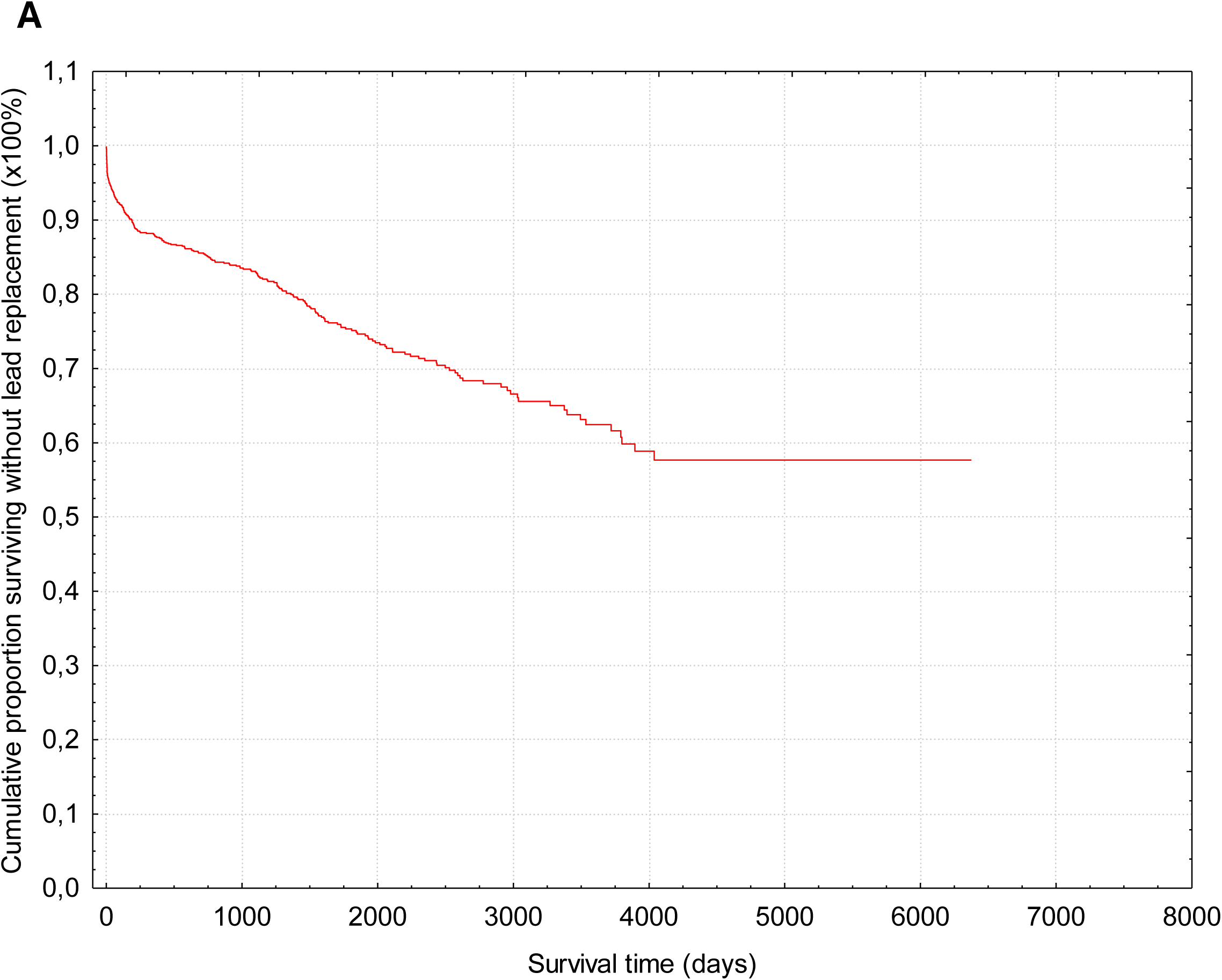
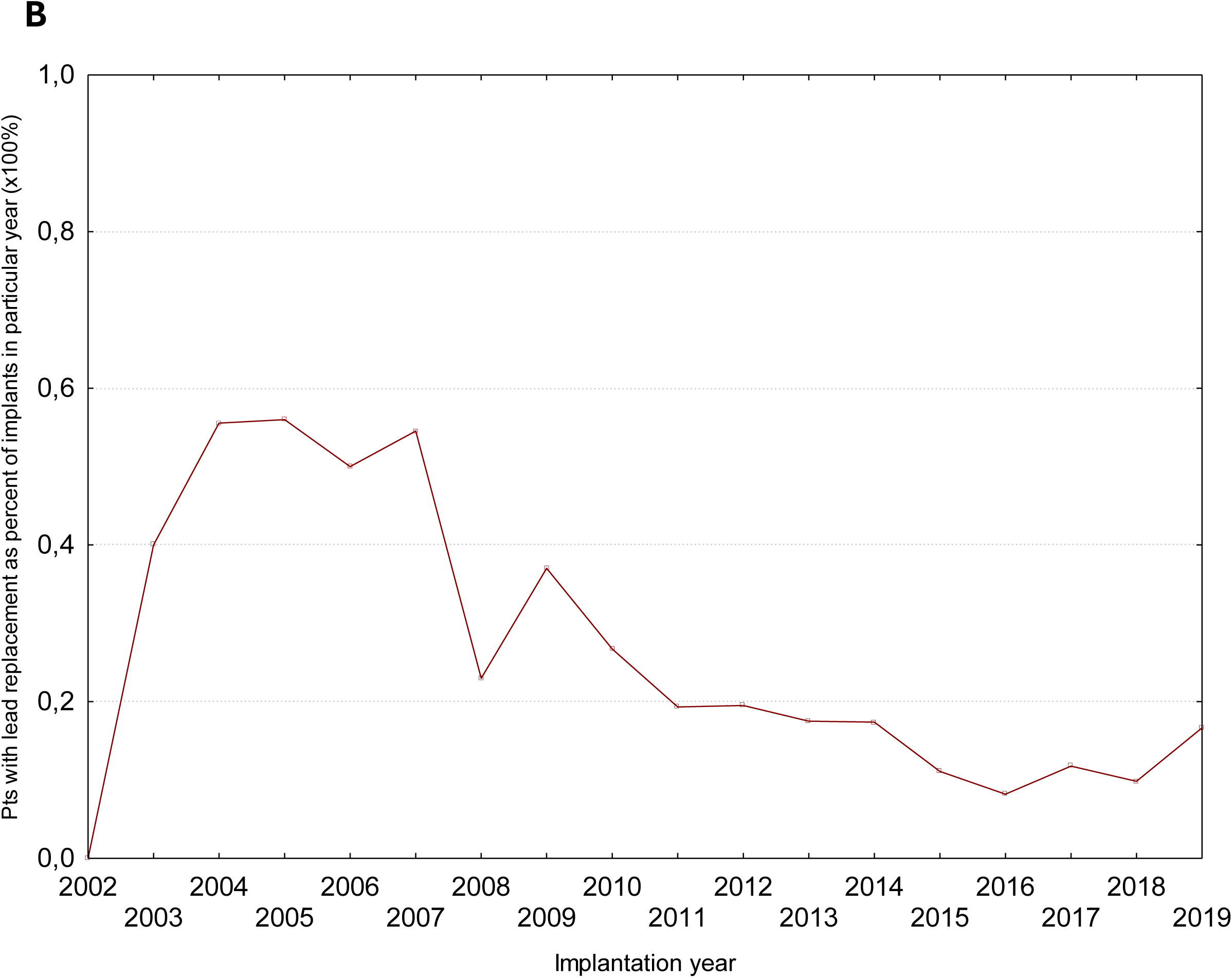
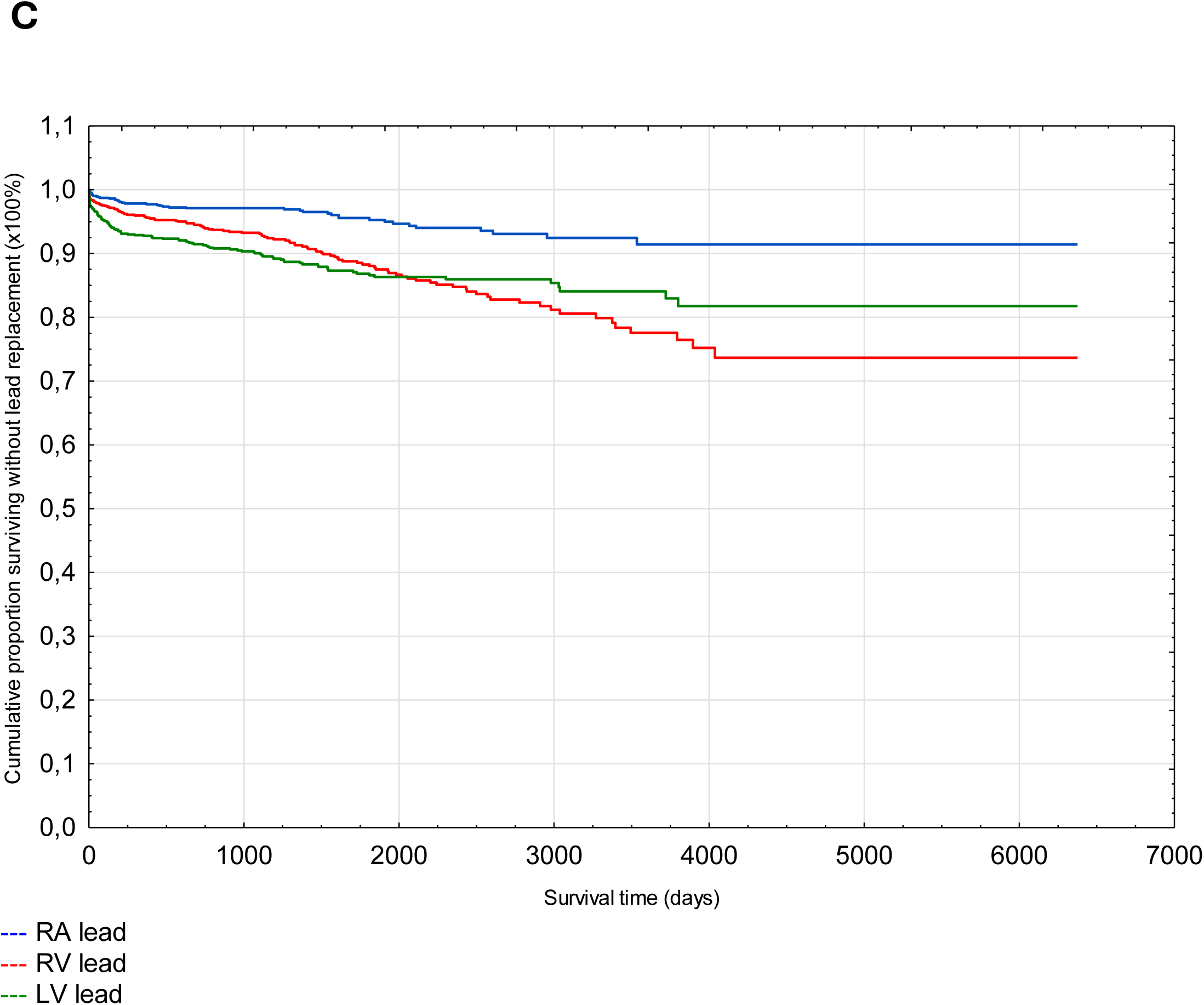
Kaplan-Meier curves of lead replacement – cumulative incidence Panel A: Cumulative incidence of lead replacement during the whole follow-up Panel B: Leads replacement during follow-up, depending on implant year Panel C: Cumulative incidence of atrial and ventricular lead replacement during the whole follow-up Abbreviations: LV – left ventricle lead, RA – right atrium lead, RV – right ventricle lead

The median time from CRT implantation to the first lead replacement was 359 days (42–1413). Patients with electrode replacement during follow-up were younger (63 vs 66 years, P<0.001), had more often dilated cardiomyopathy (45.4% vs 36.3%, P=0.009), less often ischaemic cardiomyopathy (47.4% vs 58.4%, P=0.002), permanent atrial fibrillation (12.4% vs 24.4%, P<0.001), hypertension and diabetes (52.9% vs 66.1%, P=0.002; 28.7% vs 36.5%, P=0.02), and had higher left ventricle end-diastolic diameter (68 vs 67 mm, P=0.02). The baseline characteristics of the entire population and the study groups are presented in Table 1. The duration of CRT implantation (considering *de novo* implantations) and fluoroscopy time were higher compared to patients who did not require lead replacement over time (135 vs 120 min, P<0.001, and 20.5 vs 16 min, P<0.001, respectively).

**Table 1.**
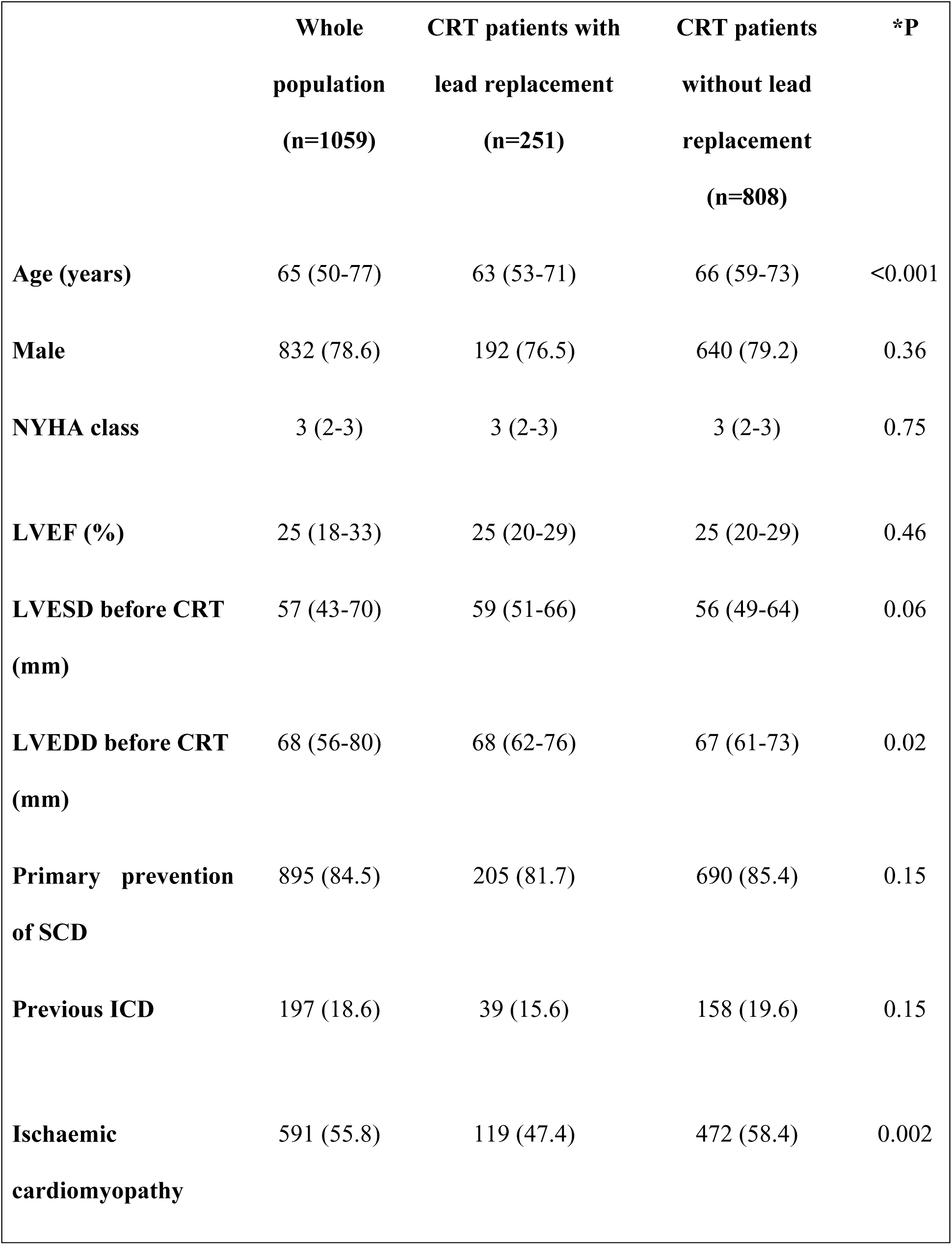
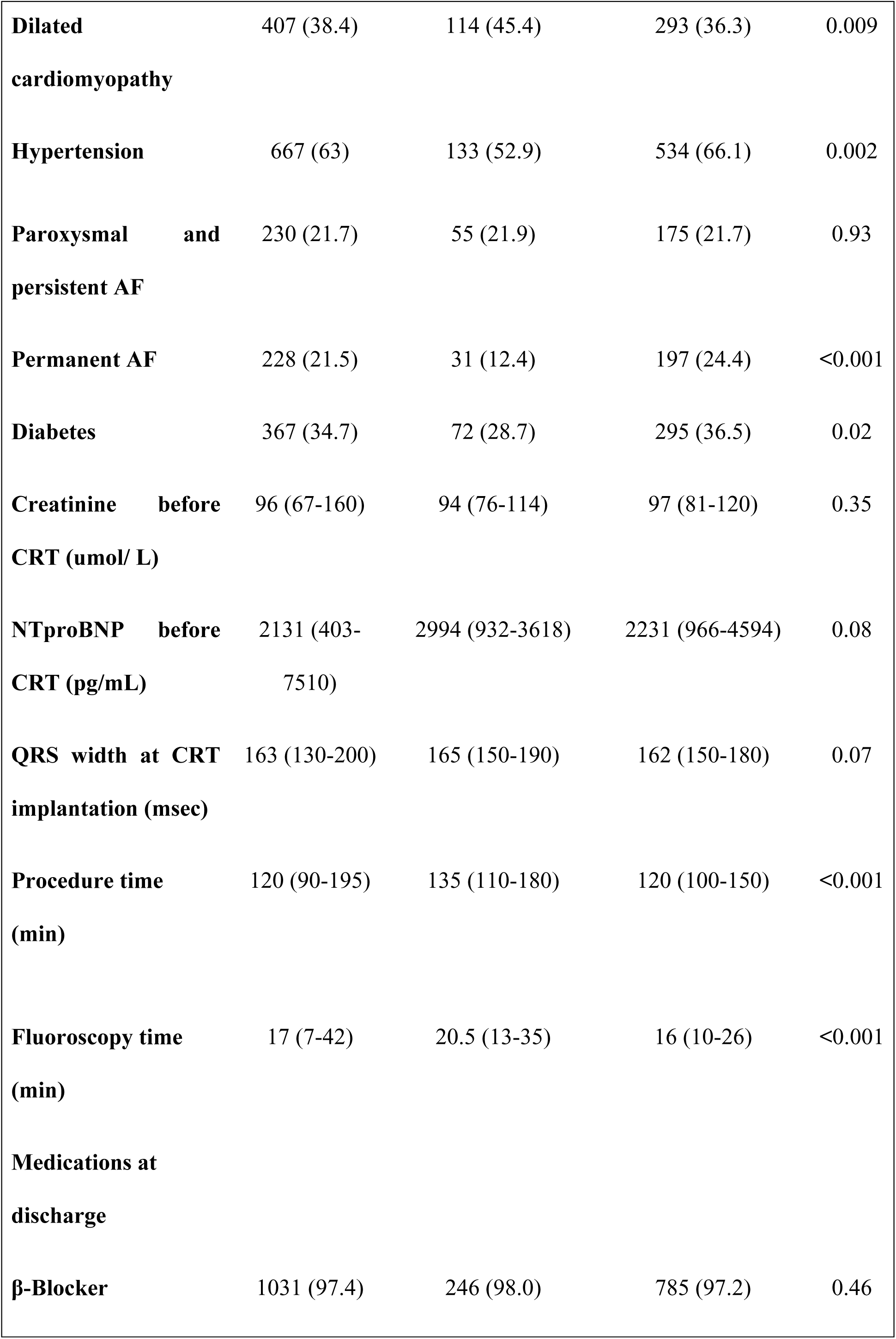
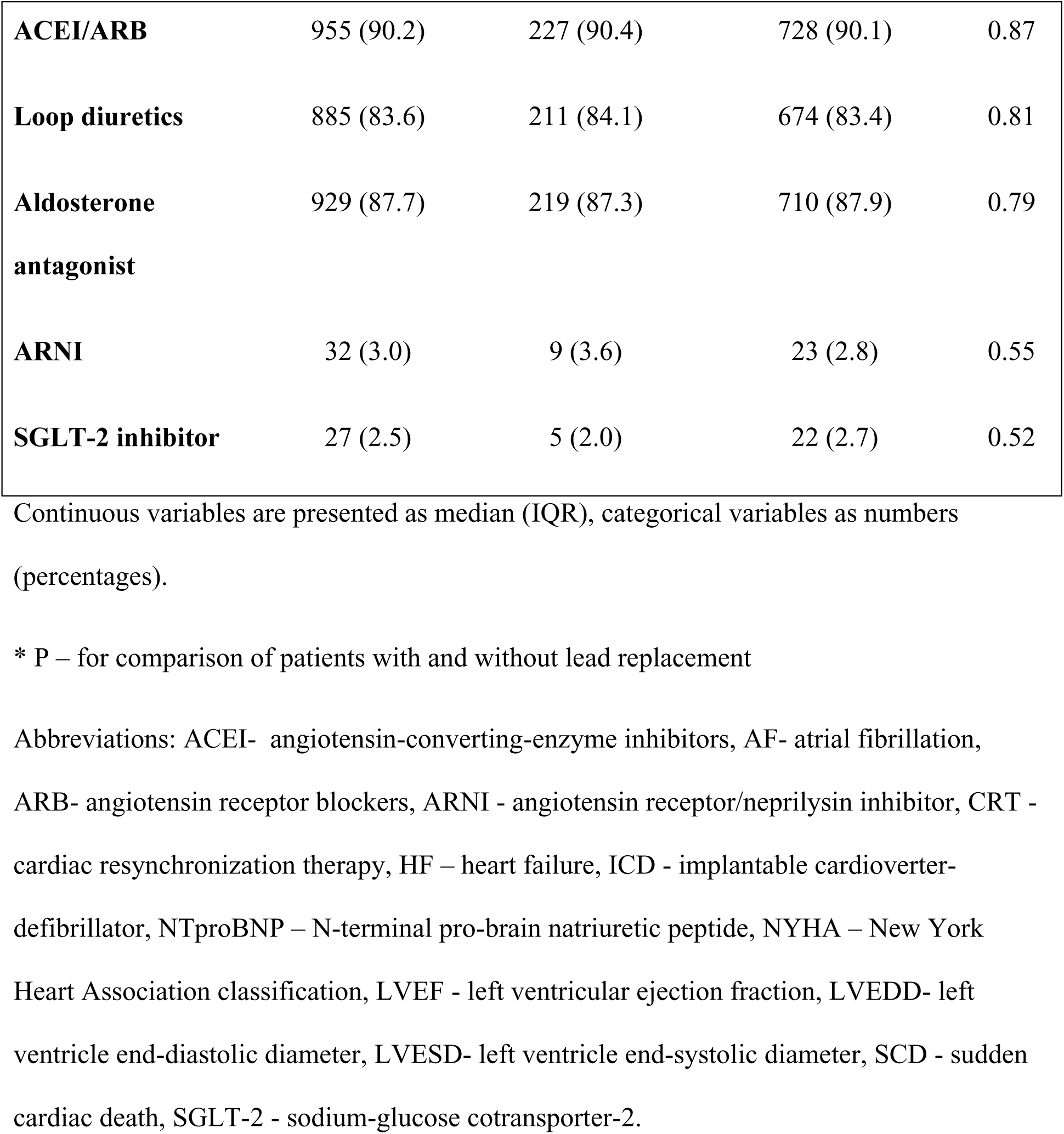
Baseline characteristics of the whole population, patients with and without lead replacement.

### Independent predictors of lead replacement during follow-up

The multivariable Cox regression model for lead replacement in patients undergoing CRT is presented in Table 2. None of the baseline patient characteristics were found to predict lead replacement independently. The index procedural data was the only factor independently associated with the need for lead replacement. In multivariable regression analysis, longer duration of the first CRT device implantation and higher fluoroscopy time were identified as independent predictors of lead replacement during FU (HR 1.03, 95% CI 1.01–1.06, P=0.02 and HR 1.11, 95% CI 1.02–1.15, P=0.01, respectively).

**Table 2.** Multivariable Cox regression model for lead replacement in patients treated with conventional CRT.

| **Variable** | **HR (± 95% CI)** | **P** |
| --- | --- | --- |
| **Age** | 0.99 (0.99-1.01) | 0.59 |
| **Procedure time (min)** | 1.03 (1.01-1.06) | 0.02 |
| **Fluoroscopy time (min)** | 1.11 (1.02-1.15) | 0.01 |
| **Ischaemic cardiomyopathy** | 1.01 (0.68-1.51) | 0.94 |
| **Dilated cardiomyopathy** | 1.17 (0.79-1.72) | 0.44 |
| **Hypertension** | 0.79 (0.61-1.04) | 0.1 |
| **Diabetes** | 0.95 (0.71-1.26) | 0.7 |
| **Permanent AF** | 0.68 (0.59-1.06) | 0.06 |
| **LVEDD before CRT** | 1.01 (0.99-1.02) | 0.16 |

Abbreviations: AF – atrial fibrillation, CI – confidence interval, CRT - cardiac resynchronization therapy, HR – hazard ratio, LVEDD - left ventricle end-diastolic diameter.

### Type and time of electrode replacement

Overall, 119 patients (11.2%) required replacement of the LV lead, 114 subjects (10.8%) needed a right ventricular (RV) lead replacement, and 42 patients (4%) required a right atrial (RA) lead replacement. Figure 1C shows the cumulative incidence of individual lead replacements. In 19 patients (1.8%), an additional subcutaneous lead was needed due to ineffective shock therapy for ventricular fibrillation.

Of those who required lead replacement, 126 subjects (50.2%) underwent a lead-related re-do intervention within the first year following CRT implantation. Within the first year after CRT implantation, a total of 135 leads were replaced: 68 LV leads (50.4%), 45 RV leads (33.3%), and 22 RA leads (16.3%).

Overall, 143 patients (13.5%) had a follow-up period of more than ten years. Of these, 67 subjects (46.8%) experienced one or more lead replacements over time. In patients with a follow-up of over 10 years, 23 (16.1%) subjects had a LV lead replacement. The relationship between replaced lead type and follow-up duration is shown in Table 3 and Figure 2.

**Figure 2.**
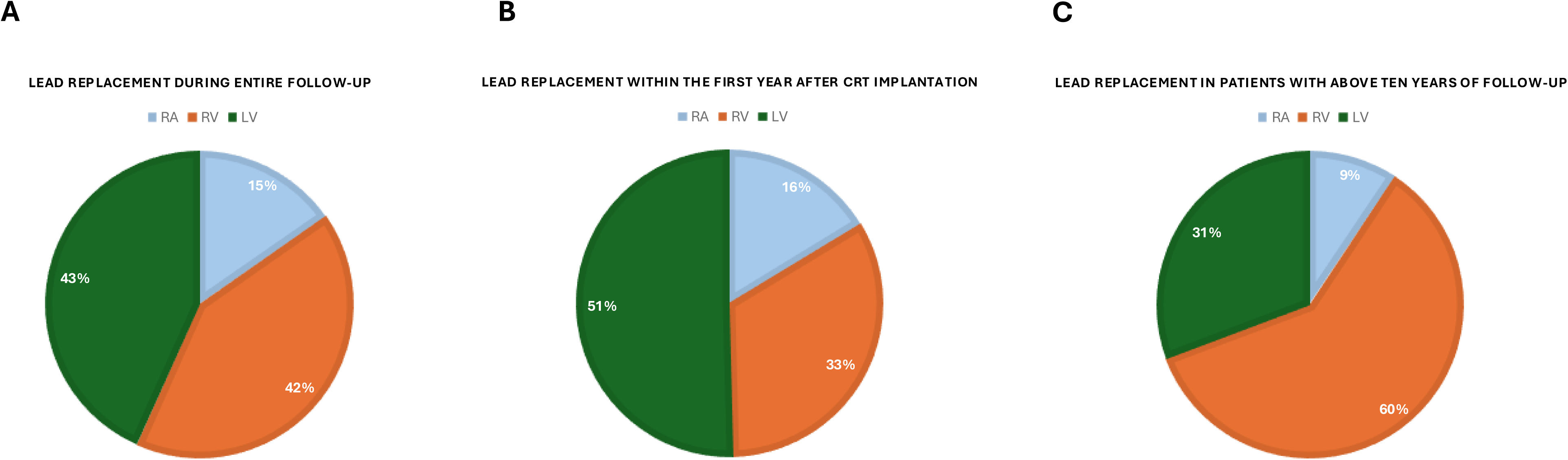
Lead replacement in CRT patients. **A. During the entire follow-up** **B. Within the first year after CRT implantation** **C. Over 10 years of follow-up** Abbreviations: LV – left ventricle lead, RA – right atrium lead, RV – right ventricle lead

**Table 3.** Patients requiring lead replacement depend on the timing of their follow-up.

| **Electrode** | **During entire follow-up (n=1059)** | **Within the first year after CRT implantation (n=1059)** | **In patients with above ten years of follow-up (n=143)** |
| --- | --- | --- | --- |
| **RA – no. (%)** | 42 (4) | 22 (2.1) | 7 (4.9) |
| **RV – no. (%)** | 114 (10.8) | 45 (4.2) | 45 (31.5) |
| **LV – no. (%)** | 119 (11.2) | 68 (6.4) | 23 (16.1) |

Abbreviations: CRT – cardiac resynchronization therapy, LV – left ventricle lead, RA – right atrium lead, RV – right ventricle lead

From 2002 through 2010, as many as 160 out of 445 patients (35.9%) required lead replacement, whereas from 2011 through 2019, 91 out of 614 subjects (14.8%) underwent lead replacement (P < 0.001). In the respective time periods, that is 2002-2010, and 2011-2019, there were 67/445 (15.1%), and 52/614 (8.5%) patients who needed LV lead replacement (P<0.001). Of note, the median follow-up duration for the time period 2002-2010 was 2413 (1061–3921) days, while for the 2011-2019 time period, it was 1330 (723–2185) days (P<0.001), respectively. In the first year after CRT implantation, 33 LV leads (33/445, 7.4%) and 35 LV leads (35/614, 5.7%; P=0.26) were replaced during the periods 2002-2010 and 2011-2019, respectively. Lead replacement during follow-up, depending on implant year, has been illustrated in Figure 1 B.

### Outcome

During the median follow-up of 1,661 days (range: 815-2,792 days), the overall all-cause mortality was 54.3% (n=575). Specifically, 128 (51%) and 447 (55.3%; P=0.3) patients died in the group with and without the need for lead replacement, respectively. Figure 3 presents Kaplan-Meier survival curves comparing patients who underwent lead replacement with those who did not.

**Figure 3.**
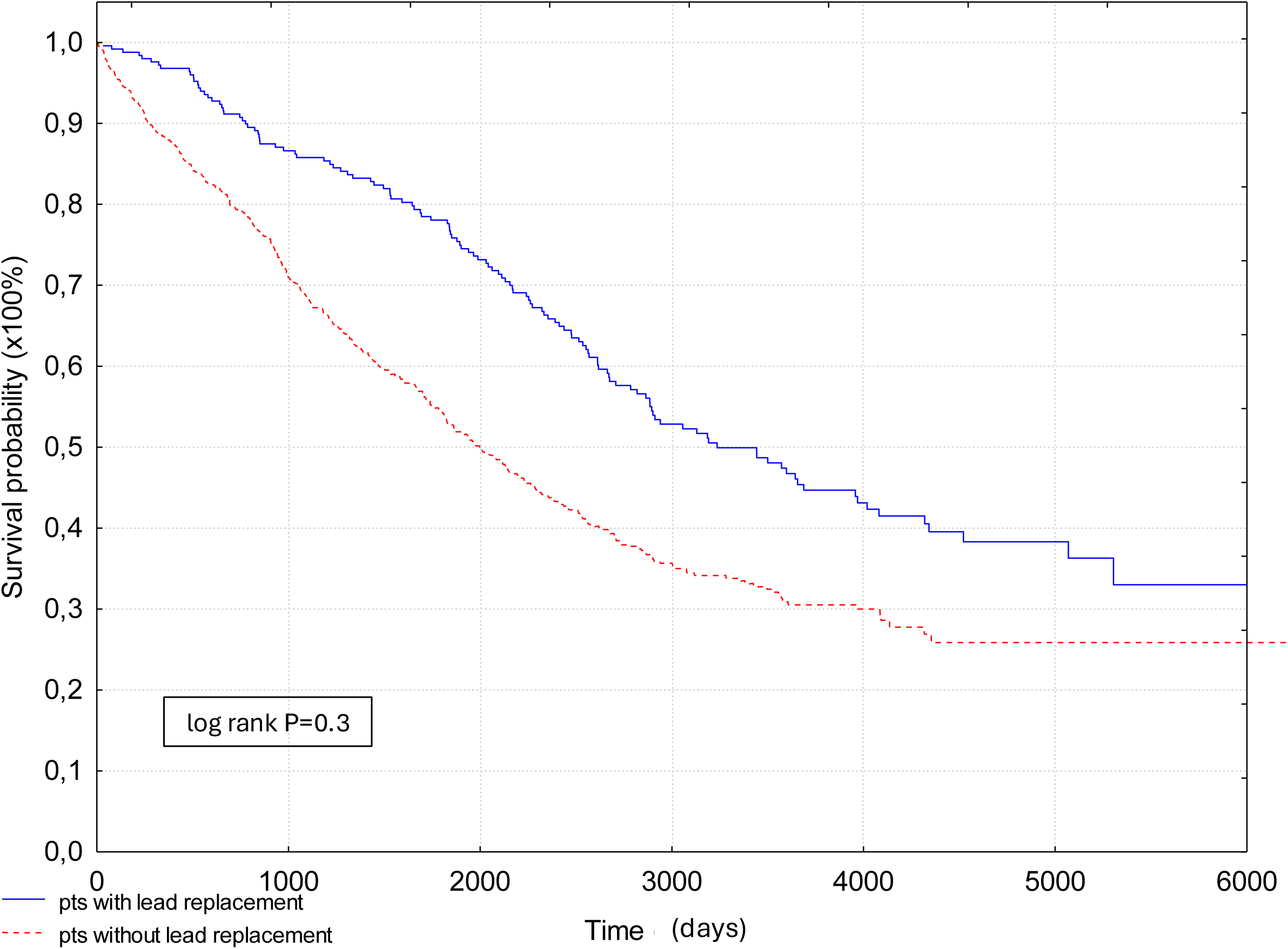
Kaplan-Meier curves of survival in patients with and without lead replacement during follow-up. Figure legend: pts – patients

Cardiac device-related infective endocarditis (CDRIE) developed in 36 (4.5%) patients who did not require lead replacement and in 14 (5.6%) patients who underwent a redo procedure and lead replacement, respectively (P = 0.82). Figure 4 presents Kaplan-Meier survival curves for patients with and without cardiac device-related infective endocarditis, comparing those with and without lead replacement.

**Figure 4.**
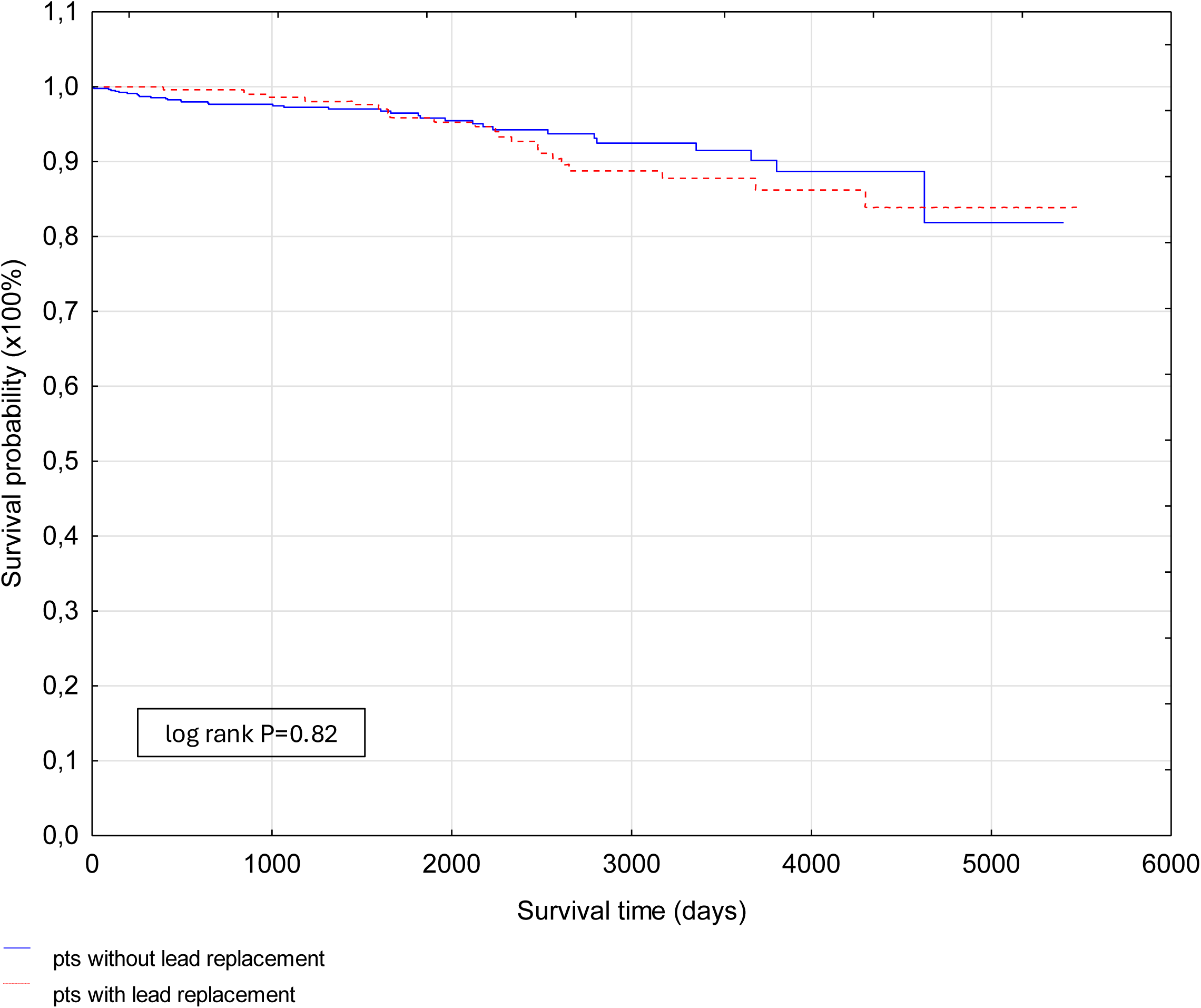
Kaplan-Meier curves of survival without cardiac device-related endocarditis in patients with and without lead replacement during follow-up. Figure legend: pts – patients

## Discussion

The main findings of our study are as follows: 1) In the observation of over ten years, almost half of all patients need one or more lead replacements; 2) In a real-world registry, about 25% of patients receiving conventional CRT require lead replacement within 4.5 years due to non-infectious reasons; 50% of those need lead replacement within the first year; 3) Overall, leads that require replamacent over time are as follows: left ventriclular (11.2%), right ventricular (10.8%) and atrial (4%) leads, respectively; 4) Lead replacement in the first year was most commonly related with left ventricular coronary sinus lead (51%), while during the long-term FU (>10 years of observations) with the defibrillation lead (61%); 5) Duration of the index procedure (CRT implantation) is the only independent predictor of lead replacement over time; 6) Re-do procedures and lead replacements do not affect long-term mortality nor the incidence of cardiac device-related infective endocarditis.

### Incidence of lead replacement in CRT patients

Several studies have assessed lead failure rates and the type of failure; however, these rates vary depending on the types of leads, duration of the follow-up period, characteristics of the studied population, type of venous access, and the definition of failure (6, 7, 8, 9, 10). One of the most critical issues is the structure of the lead. The manufacturers provide survival probability reports for their leads in adult patients. However, they are limited to one type of lead. Many published data focus on dysfunction of a specific type of lead (e.g. Sprint Fidelis [Medtronic] and Riata [Abbott]) (10, 11, 12, 13), and they do not provide data on the entire population of patients with ICD or CRT with different types of leads implanted over a long period of time and observed subsequently for many years. Pacing leads had lower failure rates than ICD because of their less complex construction (14). Furthermore, manufacturers continuously improve the quality of electrodes, which may lead to their increased long-term survival (15). There are now DF-4 instead of DF-1 connectors in ICD leads, quadripolar instead of bipolar LV leads, etc (16). Data regarding the long-term performance of LV leads is also limited. The mean follow-up duration of most published data is 2 to 3 years, and data above 10 years are very limited; however, older leads are associated with an increased failure risk. Some studies reported a 17-20% ICD lead failure rate at 10 years (6, 16). Of note, ICD lead fractures were more common in the ICD-CRT than in the ICD group (11). In our opinion, lead durability should be assessed separately for CRT and ICD, because CRT is a much more complex device, with a larger can and three leads (including a LV coronary sinus lead), which increases the risk for both lead dislocation and damage. Additionally, CRT implantation is more challenging, and the procedure is longer. Insertion of three leads close to each other, e.g., *via* a subclavian vein, further increases the risk of lead damage due to the proximity of the leads and their continuous interaction. Finally, as compared with ICD, CRT is implanted in patients with the most advanced HF and increased risk for complications *per se* (including infectious ones) due to HF itself.

The second main reason for lead replacement is lead dislodgement, which varies from 2.9% to 10.6% depending on follow-up duration, studied population, and the type of analyzed lead (17, 18). Lead failures are more frequent in ICDs than in pacemaker electrodes (14). With regard to lead dislodgment, higher rates were reported for LV leads as compared with RA or RV (6.8%, 1%, and 0.6%, respectively) (17, 18). Main reasons for that may be, for example, passive fixation of the LV compared with active fixation of right-heart leads, along with securing a stable lead position in coronary sinus branches due to challenging anatomy.

### Independent predictors of lead replacement

Data on independent predictors of lead failure are scarce. To date, the main research has focused specifically on Sprint Fidelis and Riata electrodes (11, 19, 20). Reported independent risk factors for Sprint Fidelis lead damage were higher LVEF, younger age, female gender, noncephalic venous access, and previous lead damage (11, 19, 20). In contrast to these reports, in our study, we have analyzed in detail the high-volume dataset of 1059 consecutive patients implanted with CRT who were followed for over 10 years and searched for lead failure predictors, taking into account a broad spectrum of varius important clinical factors such as patients’ clinical characteristics, procedural data, operators’ experience, lead type, concomitant medical therapy etc. Importantly, we found no correlation among all tested parameters except for procedure duration, which appeared to be the only independent risk factor for a re-do procedure and lead replacement. This finding emphasizes that prolonged CRT implantation, *per se*, due to, for example, challenging anatomy, substantially and independently increases the risk of a re-do procedure.

### Long-term prognosis and risk of CDRIE in CRT patients with lead replacement

Similarly to very limited reports on risk factors for lead replacements, data on the long-term prognosis of patients following lead replacement are also scarce (21, 22, 23).

Specifically, to the best of our knowledge, data on the prognosis of CRT patients who required *versus* did not need lead revisions/extraction/replacement have not yet been reported. Importantly, our data show that lead-related re-do procedures do not impact the long-term prognosis of CRT recipients. Namely, lead replacement neither increased the risk of all-cause mortality nor device-related endocarditis. These data may be reassuring, as CRT recipients are perceived to be at high risk of complications of any re-do procedures.

### Study limitations and strengths

A single-center study design is a limitation of our observations. A relatively small group of patients is another one, but on the other hand, the number of patients was sufficient for statistical purposes. We did not analyze the need for lead replacement with regard to initial lead location, e.g., RV lead in the apex or septal site, and the LV lead in the lateral, posterolateral vein, etc. All patients had their LV leads implanted *via* subclavian vein puncture. Many of the RA and RV leads were also implanted via this approach. Due to the lack of precise data, a detailed analysis of any correlation between vein access site and lead failure was not feasible.

Our study provides long-term follow-up data on patients with the most complex CIEDs in a real-world setting. We present our unique findings, which show that, in over 10 years of observation in our continuously updated dataset of consecutive CRT recipients, nearly every second patient requires lead replacement over time, but a re-do procedure does not deteriorate long-term prognosis.

## Conclusions

Almost 25% of patients with traditional CRT require lead replacement within 4.5 years due to non-infectious reasons. Half of the patients have a lead replacement within the first year after CRT implantation. Duration of the index procedure (CRT device implantation) and fluoroscopy time are strong predictors for lead replacement over time. Almost half of all patients with a follow-up period of over ten years have one or more lead replacements. Patients with and without lead replacement have similar mortality.

## Data Availability

none

## Funding sources

None

## Conflict of interest

E. Jędrzejczyk-Patej, M. Mazurek, O. Kowalski, A. Sokal - consultant fees from Medtronic, Biotronik, Abbott and Boston Scientific; R. Lenarczyk – reports funding from the European Union’s Horizon 2020 research and innovation programme under grant agreement no 847999; Z. Kalarus – speaker bureaus for Bayer, BMS/Pfizer, Boehringer-Ingelheim, Elli-Lilly, Abbott; other authors – no conflict of interests reported.

## Abbreviations

CRT: cardiac resynchronization therapy
HF: heart failure
LVEF: left ventricle ejection fraction
NYHA: New York Heart Association

## Notes

### Competing Interest Statement

E. Jedrzejczyk-Patej, M. Mazurek, O. Kowalski, A. Sokal - consultant fees from Medtronic, Biotronik, Abbott and Boston Scientific; R. Lenarczyk reports funding from the European Union's Horizon 2020 research and innovation programme under grant agreement no 847999; Z. Kalarus speaker bureaus for Bayer, BMS/Pfizer, Boehringer-Ingelheim, Elli-Lilly, Abbott; other authors no conflict of interests reported.

### Author Declarations

The study protocol has been approved by the Ethics Committee of the Medical University of Silesia (the Institutional Review Board [IRB] number assigned: BNW/NWN/0052/KB/30/24).

